# Aficamten in Patients With Obstructive Hypertrophic Cardiomyopathy: An Integrated Safety Analysis

**DOI:** 10.64898/2026.03.06.26347726

**Authors:** Ahmad Masri, Martin S. Maron, Roberto Barriales-Villa, Robert M. Cooper, Perry Elliott, Michael A. Fifer, Pablo Garcia-Pavia, Anjali T. Owens, Scott D. Solomon, Albree Tower-Rader, Camelia Dumitrescu, Justin Godown, Stephen B. Heitner, Daniel L. Jacoby, Stuart Kupfer, Fady I. Malik, Regina Sohn, Jenny Wei, Sara Saberi, REDWOOD-HCM, SEQUOIA-HCM, MAPLE-HCM, and FOREST-HCM Investigators

## Abstract

**BACKGROUND:** Aficamten is a next-in-class, oral, selective cardiac myosin inhibitor approved for the treatment of obstructive hypertrophic cardiomyopathy (oHCM). A comprehensive understanding of long-term safety is essential to inform clinical use.

**OBJECTIVE:** To assess the integrated safety profile of aficamten across phase 2/3 clinical trials in patients with oHCM.

**METHODS:** This integrated safety analysis pooled data from patients with oHCM who received ≥1 dose of aficamten or placebo/metoprolol in REDWOOD-HCM, SEQUOIA-HCM, MAPLE-HCM, and FOREST-HCM. Safety outcomes included treatment-emergent adverse events (TEAEs), serious TEAEs, adverse events of special interest, occurrences of site-read left ventricular ejection fraction (LVEF) <50%, and echocardiography-guided treatment modifications. Events were summarized descriptively and using exposure-adjusted incidence rates (EAIRs) per 100-patient-years.

**RESULTS:** The cumulative aficamten-treated pool included 463 unique patients, representing 697 patient-years of exposure. Aficamten was well tolerated, with permanent treatment discontinuation occurring in 4 (0.9%) aficamten-treated patients (EAIR 0.6). In the control group pool, rates of TEAEs were comparable between aficamten and placebo/metoprolol, except hypertension was more common in aficamten-treated patients. In the cumulative aficamten-treated pool, LVEF <50% occurred in 19 (4.1%) patients (EAIR 2.8). There were no cases of LVEF <50% associated with clinical heart failure that were attributable to aficamten, and no excursions of LVEF <40%. New-onset atrial fibrillation was uncommon (EAIR 2.4).

**CONCLUSIONS:** Over nearly 700 patient-years of exposure, aficamten was well tolerated with a favorable safety profile in patients with oHCM. The rates of clinically relevant systolic dysfunction, atrial fibrillation, and other major cardiovascular events were low and similar to placebo or metoprolol.

**Clinical trial registration:** REDWOOD-HCM (<u>NCT04219826</u>); SEQUOIA-HCM (<u>NCT05186818</u>); MAPLE-HCM (<u>NCT05767346</u>); FOREST-HCM (<u>NCT04848506</u>)

## INTRODUCTION

Hypertrophic cardiomyopathy (HCM) is a genetic disease of the myocardium characterized by sarcomeric hypercontractility, left ventricular hypertrophy, and impaired myocardial relaxation.^1^ Excessive actin-myosin cross-bridge formation leads to increased contractile force, elevated intracavitary pressures, and progressive symptoms including exertional dyspnea, chest pain, syncope, and heart failure across the spectrum of HCM phenotypes.^2,3^ In many patients, these abnormalities are accompanied by dynamic left ventricular outflow tract (LVOT) obstruction, defining obstructive HCM (oHCM). Traditional pharmacologic therapies for oHCM, such as beta-blockers, non-dihydropyridine calcium-channel blockers, and disopyramide, are limited by modest efficacy and poor tolerability in many patients.^4^

Cardiac myosin inhibitors represent a targeted therapeutic approach that directly addresses the underlying pathophysiology of HCM. Aficamten is a next-in-class, oral, selective cardiac myosin inhibitor that was recently approved by the United States Food and Drug Administration (FDA) for the treatment of symptomatic oHCM.^5^ Aficamten reduces myocardial hypercontractility by stabilizing myosin in a low-energy, pre-power stroke state, thereby decreasing excessive actin–myosin interactions.^6^ Through this mechanism, aficamten improves functional capacity, reduces LVOT gradients, and improves symptoms in patients with oHCM.^7^ However, because cardiac myosin inhibitors, including aficamten, act directly at the level of the cardiac sarcomere to reduce myocardial contractility, there is a risk for unintended excessive reductions in left ventricular ejection fraction (LVEF) and the development of heart failure. Accordingly, both aficamten and mavacamten (first-in-class myosin inhibitor) are approved with their molecule-specific risk evaluation and mitigation strategy (REMS) programs with elements to help ensure safe use primarily through periodic monitoring with echocardiograms.^8,9^ Despite a shared therapeutic class, aficamten and mavacamten differ in key pharmacologic properties, including target binding site, half-life, metabolic pathways, and exposure-response characteristics.^10^ Understanding the implications of these differences requires a comprehensive assessment of cumulative safety data across clinical trials with varying durations of exposure, which can inform real-world clinical practice.

To date, aficamten has been studied across multiple clinical trials, including randomized placebo-controlled studies, an active-comparator trial (vs metoprolol), and an ongoing long-term open-label extension. While individual trials have demonstrated favorable efficacy, safety, and tolerability,^11,12^ this analysis, which integrates safety data across all available aficamten studies, is key for developing an understanding of the frequencies and rates of adverse events, particularly in the setting of a relatively rare disease such as oHCM. As such, the objective of this study is to evaluate the integrated safety profile of aficamten in patients with oHCM by leveraging cumulative data from all completed and ongoing clinical trials, with a particular focus on adverse events related to decreased LVEF.

## METHODS

### STUDY DESIGN AND DATA SOURCES

This integrated safety analysis included data from four clinical trials evaluating aficamten in patients with oHCM: (1) REDWOOD-HCM (Randomized Evaluation of Dosing With CK 3773274 in Obstructive Hypertrophic Cardiomyopathy; NCT04219826), (2) SEQUOIA-HCM (Safety and Efficacy of CK 3773274 in Adults With Symptomatic Obstructive Hypertrophic Cardiomyopathy; NCT05186818), (3) MAPLE-HCM (Metoprolol Versus Aficamten in Patients With Obstructive Hypertrophic Cardiomyopathy; NCT05767346), and (4) FOREST-HCM (Follow Up Open Label Research Evaluation of Sustained Treatment With Aficamten in Hypertrophic Cardiomyopathy; NCT04848506). The design and primary results of these studies have been reported previously,^4,7,13,14^ and key trial characteristics are summarized in Supplemental Table 1. The data cutoff date for this integrated safety analysis was May 9, 2025.

### STUDY OVERSIGHT

All trials were conducted in accordance with the principles of the Declaration of Helsinki and Good Clinical Practice guidelines. Study protocols were approved by institutional review boards or ethics committees at each participating site, and all patients provided written informed consent prior to enrollment. Independent data monitoring committees oversaw patient safety in the blinded studies.

### STUDY POPULATIONS

Two study populations were defined: (1) the cumulative aficamten-treated pool included all patients with oHCM who received at least 1 dose of aficamten in REDWOOD-HCM, SEQUOIA-HCM, MAPLE-HCM, or FOREST-HCM; and (2) the control group pool, which included patients randomized to placebo in REDWOOD-HCM and SEQUOIA-HCM or to metoprolol in MAPLE-HCM. For contextual comparisons, patients randomized to aficamten in the controlled studies (REDWOOD-HCM cohorts 1 and 2, SEQUOIA-HCM, and MAPLE-HCM) were analyzed independently to allow direct comparison with the control group pool. Subsequently, all patients who received at least 1 dose of aficamten across both controlled and open-label studies were analyzed together as the cumulative aficamten-treated pool to characterize longer-term safety across the full exposure period.

### TREATMENT EXPOSURE AND DOSE DEFINITIONS

Aficamten was administered using study-specific titration algorithms designed to achieve therapeutic reduction in LVOT gradients while maintaining LVEF ≥50% (Supplemental Table 2). Maintenance dose was defined as the dose achieved after completion of titration. The maintenance phase was defined as the period beginning at the maintenance dose visit and continuing through subsequent follow-up. Total exposure was summarized as duration of treatment in months and cumulative patient-years of aficamten exposure.

### SAFETY ASSESSMENTS

Safety evaluations included treatment-emergent adverse events (TEAE), adverse events of special interest, and treatment interruptions or discontinuations. TEAEs were defined as adverse events occurring after the first dose of study drug through the end of follow-up. Adverse events were coded using the Medical Dictionary for Regulatory Activities, and drug relatedness and severity was assessed by site investigators. Adverse events of special interest were prespecified and included LVEF excursions <50%, atrial fibrillation (classified as either new onset or recurrent), heart failure, syncope, stroke, myocardial infarction, and cardiovascular death. Classification of atrial fibrillation as new onset or recurrent was based on the presence or absence of a documented history of atrial fibrillation in baseline medical history.

LVEF was assessed in real-time by transthoracic echocardiography, and site readings served as the basis for echocardiographic analyses for this study. Events of LVEF <50% were evaluated in relation to subsequent clinical management, including dose down-titration, treatment interruption, or permanent discontinuation. A management decision was attributed to a low LVEF event when it occurred within ±7 days of an LVEF <50%. The clinical utility of echocardiographic monitoring during the maintenance phase was evaluated by quantifying the total number of echocardiograms performed and the proportion that resulted in treatment modifications.

### STATISTICAL ANALYSIS

No formal hypothesis testing was performed, and all analyses were descriptive. Continuous variables were summarized using means and SDs or medians and interquartile ranges, as appropriate. Categorical variables were summarized using counts and percentages. Event rates were reported as exposure-adjusted incidence rates (EAIRs) expressed as patients per 100 patient-years of exposure. Safety outcomes were summarized separately for the cumulative aficamten-treated pool and the control group pool. All analyses were performed using SAS Enterprise Guide version 8.3.

## RESULTS

### STUDY POPULATIONS AND EXPOSURE

The cumulative aficamten-treated pool included 463 unique patients with oHCM who received at least 1 dose of aficamten across REDWOOD-HCM, SEQUOIA-HCM, and MAPLE-HCM, with follow-up in FOREST-HCM, representing 697 cumulative patient-years of exposure (Figure 1). The control group pool included 240 patients randomized to placebo (n = 153) or metoprolol (n = 87) in REDWOOD-HCM, SEQUOIA-HCM, and MAPLE-HCM, which were compared to 258 patients randomized to aficamten in those studies.

**FIGURE 1.**
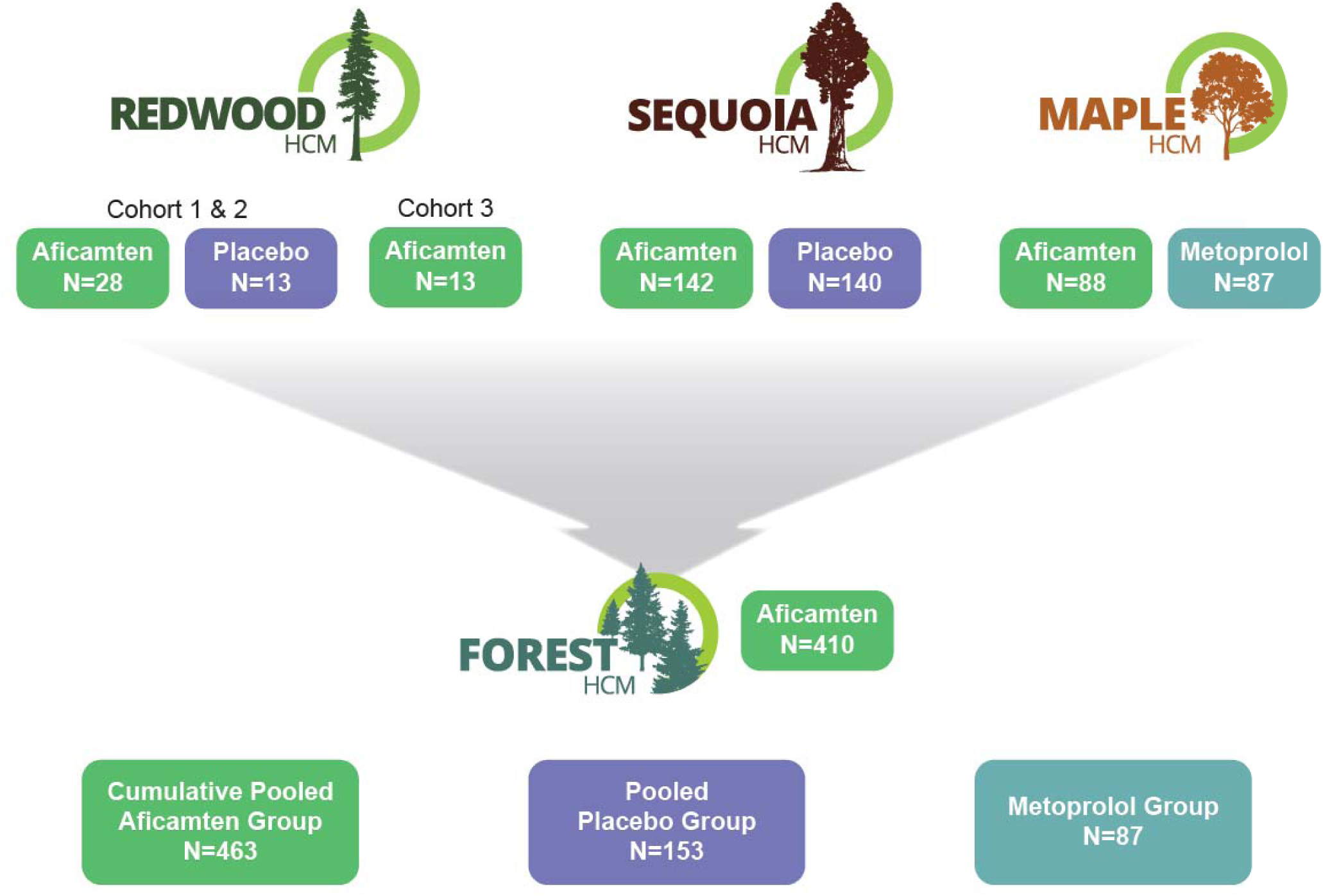
Clinical Development Program and Integrated Safety Analysis Populations in Obstructive Hypertrophic Cardiomyopathy (oHCM) REDWOOD-HCM (Cohorts 1 & 2), SEQUOIA-HCM, and MAPLE-HCM were randomized parent trials of aficamten versus placebo or metoprolol in oHCM, with eligible patients enrolling in the FOREST-HCM open-label extension. The controlled group pool included patients treated during the blinded periods (aficamten n=258, placebo n=153, metoprolol n=87). The cumulative aficamten pool comprised all patients exposed to aficamten across parent and extension studies (n=463)

Baseline demographic and clinical characteristics (Table 1) were similar across all studies and treatment groups. In the cumulative aficamten-treated pool, the mean (SD) age was 58.8 (±12.8) years, 195 (42%) were female, and 399 (86.2%) were White. At baseline, 218 (47.1%) patients were receiving background beta-blocker therapy, and the majority were assessed as being in New York Heart Association (NYHA) functional class II (n = 319 [68.9%]) or III (n = 139 [30.0%]). The mean (SD) duration of aficamten exposure was 18.1 (±12) months in the cumulative pool, and 312 (75%) patients achieved a dose of aficamten ≥15 mg daily during the maintenance phase.

**TABLE 1.**
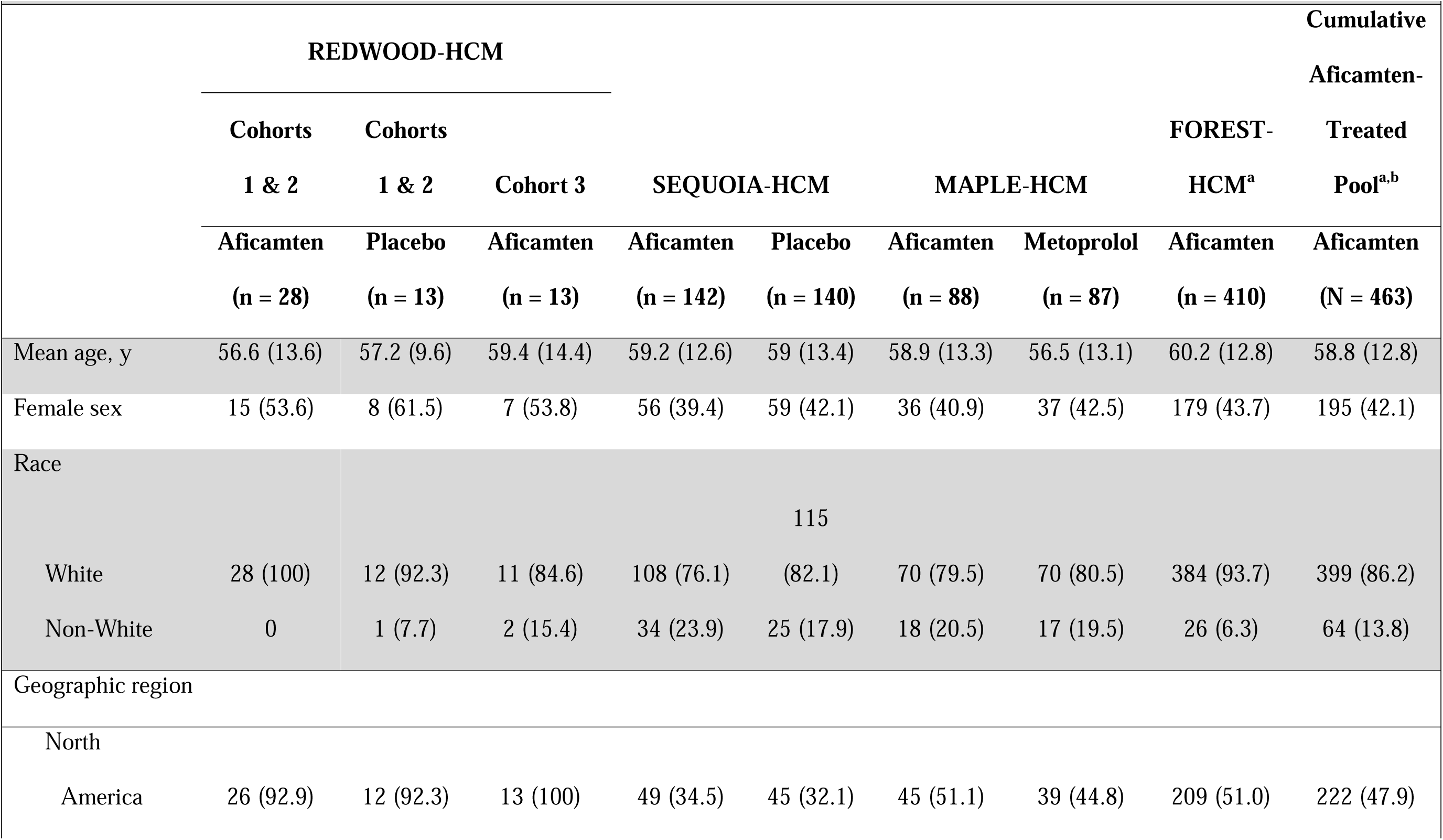
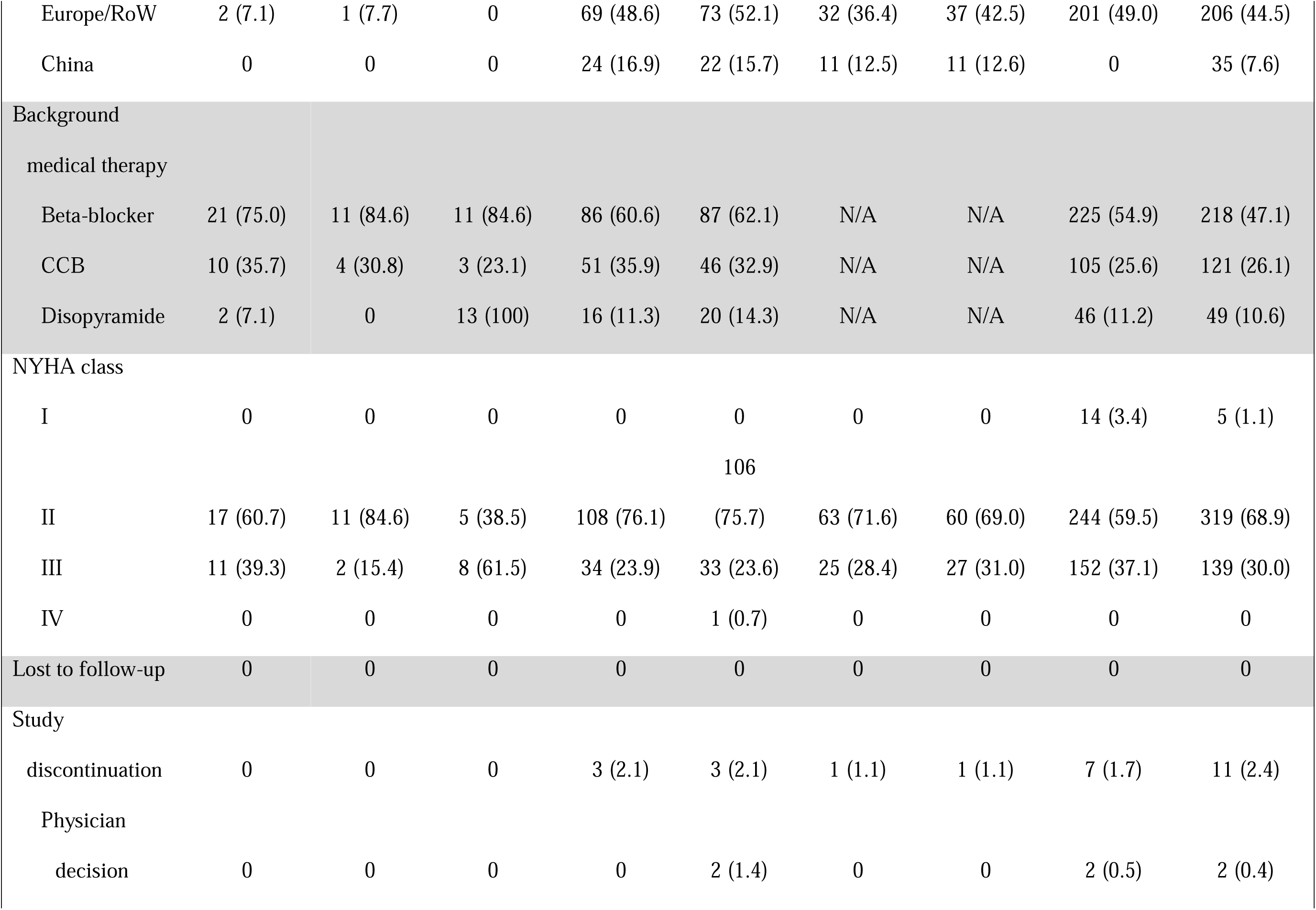
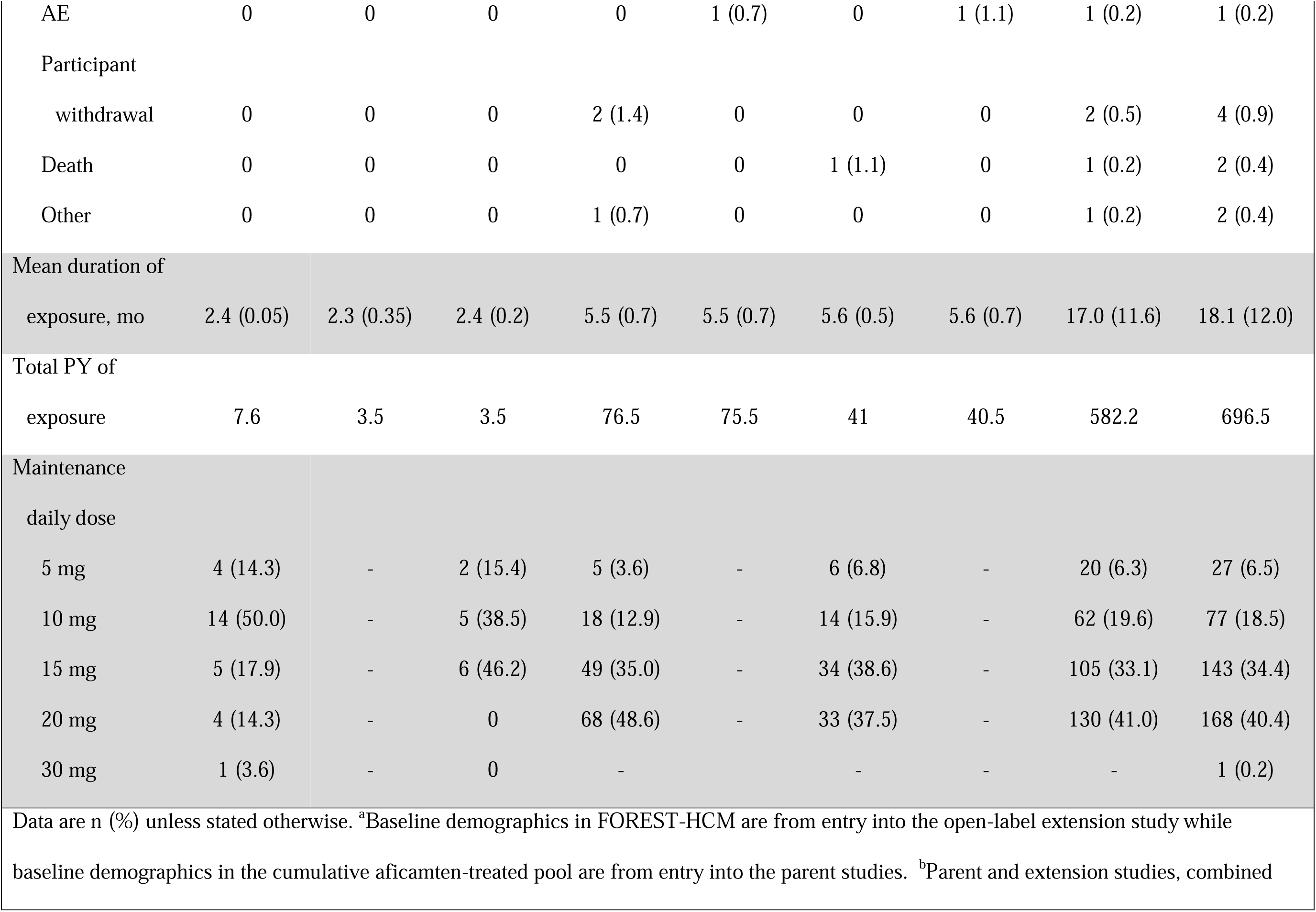
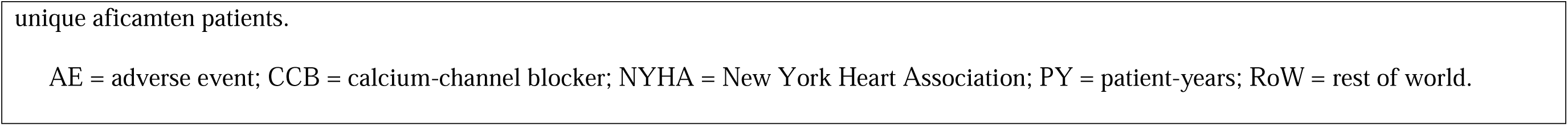
Baseline Demographics.

### OVERALL SAFETY AND ADVERSE EVENTS

A summary of adverse events is presented in Table 2. In the control group pool, the overall incidence of TEAEs was similar among patients receiving aficamten, placebo, or metoprolol. Serious TEAEs occurred in 6.6% of patients treated with aficamten compared with 9.2% in the placebo group and 6.9% in the metoprolol group. Adverse events leading to death were rare (1 [0.4%] in aficamten group and none in the placebo or metoprolol groups). TEAEs leading to drug interruption were reported in 3 (1.2%) patients in the aficamten group, 2 (1.3%) in the placebo group, and 1 (1.1%) in the metoprolol group, while permanent discontinuation occurred in 1 (0.4%), 2 (1.3%), and 3 (3.4%) patients, respectively. For adverse events occurring in ≥5% of patients, rates were similar between aficamten and comparator groups, except for hypertension. Hypertension was reported more frequently in patients treated with aficamten (n = 20 [7.8%]) compared with placebo (n = 4 [2.6%]) or metoprolol (n = 3 [3.4%]). The mean (SD) change in systolic blood pressure from baseline to the end of treatment in the controlled cohort was 3.5 (±13.6) mmHg in those randomized to aficamten compared to –2.6 (±14.1) and –5.7 (±14.5) mmHg for placebo and metoprolol, respectively.

**TABLE 2.** Overall Safety and TEAEs.

|  | From Controlled Studies<sup>a</sup> |  |  |  |  |  | Cumulative Aficamten-Treated Group |  |
| --- | --- | --- | --- | --- | --- | --- | --- | --- |
|  | Aficamten | EAIR | Placebo | EAIR |  | EAIR |  |  |
|  | ± SOC | per 100 | ± SOC | per 100 | Metoprolol | per 100 | Aficamten ± | per 100 |
|  | (n = 258) | PY | (n = 153) | PY | (n = 87) | PY | SOC (n = 463) | PY |
| TEAE |  |  |  |  |  |  |  |  |
| Serious | 17 (6.6) | 13.3 | 14 (9.2) | 18.5 | 6 (6.9) | 13.1 | 65 (14.0) | 10.1 |
| Leading to death | 1 (0.4) | 0.8 | 0 | - | 0 | - | 2 (0.4) |  |
| Leading to drug interruption | 3 (1.2) | 2.3 | 2 (1.3) | 2.6 | 1 (1.1) | 2.2 | 14 (3.0) |  |
| Leading to permanent discontinuation | 1 (0.4) | 0.8 | 2 (1.3) | 2.6 | 3 (3.4) | 6.4 | 4 (0.9) |  |
| Common TEAE ≥5% |  |  |  |  |  |  |  |  |
| COVID-19 | 16 (6.2) | 12.8 | 10 (6.5) | 13.2 | 4 (4.6) | 8.7 | 63 (13.6) | 10 |
| Upper respiratory tract infection | 21 (8.1) | 16.9 | 12 (7.8) | 16.1 | 10 (11.5) | 22.8 | 51 (11.1) | 7.6 |
| Hypertension | 20 (7.8) | 16.1 | 4 (2.6) | 5.2 | 3 (3.4) | 6.5 | 49 (10.6) | 7.4 |
| Headache | 21 (8.1) | 17.1 | 14 (9.2) | 19.1 | 6 (6.9) | 13.5 | 48 (10.4) | 7.5 |
| Dizziness | 17 (6.6) | 13.5 | 3 (2.0) | 3.9 | 15 (7.2) | 37.5 | 43 (9.3) | 6.5 |
| Dyspnea | 18 (7.0) | 14.1 | 8 (5.2) | 10.6 | 6 (6.9) | 13.4 | 41 (8.9) | 6.1 |
| Palpitations | 13 (5.0) | 10.3 | 5 (3.3) | 6.5 | 8 (9.2) | 17.7 | 38 (8.2) | 5.6 |
| Nasopharyngitis | 7 (2.7) | 5.4 | 6 (3.9) | 7.9 | 6 (6.9) | 13.1 | 34 (7.3) | 5.1 |
| Atrial fibrillation | 8 (3.1) | 6.3 | 5 (3.3) | 6.5 | 3 (3.4) | 6.4 | 29 (6.3) | 4.3 |
| Back pain | 7 (2.7) | 5.5 | 2 (1.3) | 2.6 | 2 (2.3) | 4.3 | 28 (6.0) | 4 |
| Influenza | 8 (3.1) | 6.2 | 4 (2.6) | 5.2 | 1 (1.1) | 2.1 | 27 (5.8) | 3.9 |
| Fall | 2 (0.8) | 1.5 | 3 (2.0) | 3.9 | 1 (1.1) | 2.1 | 27 (5.8) | 3.9 |
| Urinary tract infection | 6 (2.3) | 4.7 | 1 (0.7) | 1.3 | 0 | - | 26 (5.6) | 3.8 |
| Angina | 12 (4.7) | 9.4 | 6 (3.9) | 7.8 | 4 (4.6) | 8.7 | 25 (5.4) | 3.6 |
| Arthralgia | 1 (0.4) | 0.8 | 2 (1.3) | 2.6 | 2 (2.3) | 4.3 | 25 (5.4) | 3.7 |
| Sinusitis | 5 (1.9) | 3.9 | 1 (0.7) | 1.3 | 1 (1.1) | 2.1 | 24 (5.2) | 3.5 |
| AE of special interest |  |  |  |  |  |  |  |  |
| LVEF <50%<sup>b</sup> | 12 (4.7) | 9.4 | 1 (0.7) | 1.3 | 0 | - | 19 (4.1) | 2.8 |
| LVEF <50% associated with |  |  |  |  |  |  |  |  |
| non-serious AEs<sup>c</sup> | 1 (0.4) | 0.8 | 1 (0.7) | 1.3 | 0 | - | 3 (0.6) | 0.4 |
| Atrial fibrillation |  |  |  |  |  |  |  |  |
| New onset | 5 (1.9) | 3.9 | 3 (2.0) | 3.9 | 3 (3.4) | 6.4 | 17 (3.7) | 2.4 |
| Recurrent | 3 (1.2) | 2.3 | 2 (1.3) | 2.6 | 0 | - | 12 (2.6) | 1.7 |
| Heart failure | 5 (1.9) | 3.9 | 2 (1.3) | 2.6 | 1 (1.1) | 2.1 | 12 (2.6) | 1.7 |
| Stroke | 1 (0.4) | 0.8 | 1 (0.7) | 1.3 | 1 (1.1) | 2.1 | 7 (1.5) | 1 |
| Myocardial infarction | 6 (2.3) | 4.7 | 5 (3.3) | 6.5 | 4 (4.6) | 8.7 | 15 (3.2) | 2.2 |
| Syncope | 4 (1.6) | 3.1 | 3 (2.0) | 3.9 | 3 (3.4) | 6.5 | 10 (2.2) | 1.4 |
| CV death | 1 (0.4) | 0.8 | 0 | - | 0 | - | 2 (0.4) | 0.3 |

<sup>a</sup>Controlled studies include REDWOOD-HCM Cohorts 1-3, SEQUOIA-HCM, and MAPLE-HCM. <sup>b</sup>Site-read. <sup>c</sup>Included peripheral edema and dyspnea.
AE = adverse event; CV = cardiovascular; EAIR = exposure-adjusted incidence rate; LVEF = left ventricular ejection fraction; PY = patient-years; SOC = standard of care; TEAE = treatment-emergent adverse event.

Across the cumulative aficamten-treated pool, which included 463 patients and 697 patient-years of exposure, serious adverse events occurred in 14.0% of patients, corresponding to an EAIR of 10.1 per 100 patient-years. Permanent discontinuation due to adverse events was uncommon, occurring in 4 (0.9%) patients. The most frequently reported TEAEs included COVID-19, upper respiratory tract infection, hypertension, headache, dizziness, and dyspnea. EAIRs of the most common TEAEs are presented in Table 2, and the rates observed in the cumulative aficamten-treated pool were similar to or lower than those observed in the controlled studies.

### REDUCED LVEF EVENTS

A total of 19 (4.1%) patients in the cumulative aficamten-treated pool experienced at least 1 site-read LVEF <50% (EAIR 2.8 per 100 patient-years). Of these, 3 (0.6%) patients had a site-read LVEF <50% that was temporally (≤7 days) associated with non-serious isolated adverse events (peripheral edema, dyspnea, or cardiac failure); however, none were associated with reports of clinical heart failure attributed to aficamten by the investigator The report of cardiac failure occurred in a patient who experienced recurrent LVEF <50% during the parent study while receiving placebo and whose LVEF did not recover to >50% after discontinuation of aficamten in the open-label extension.

During the maintenance phase, 3,618 site-read echocardiograms were performed (mean 7.8 echocardiograms per patient). Among these, 28 (<1%) echocardiograms in 19 patients prompted dose reduction due to a site-read LVEF <50%. All but 1 of these cases demonstrated LVEF ≥50% following dose adjustment, as described above. There were no occurrences of LVEF <40%.

### ATRIAL FIBRILLATION AND OTHER EVENTS OF SPECIAL INTEREST

In controlled studies, new-onset atrial fibrillation occurred in 5 of 258 patients (1.9%; EAIR 3.9 per 100 patient-years) on aficamten compared with 3 of 153 (2.0%; EAIR 3.9 per 100 patient-years) on placebo and 3 of 87 (3.4%; EAIR 6.4 per 100 patient-years) on metoprolol. In the pooled aficamten-treated population, new-onset atrial fibrillation occurred in 17 of 463 patients (3.7%; EAIR 2.4 per 100 patient-years) (Table 2). Rates of heart failure events, syncope, stroke, myocardial infarction, and death were also low and similar between aficamten and comparator groups. There were 2 (0.4%) deaths in the cumulative aficamten-treated pool (EAIR 0.1 per 100 patient-years); however, neither was considered related to the study drug.

## DISCUSSION

In this integrated safety analysis representing nearly 700 patient-years of exposure, aficamten demonstrated a favorable safety profile in patients with symptomatic oHCM that was comparable to placebo or metoprolol. Across randomized placebo-controlled trials, an active comparator study, and a long-term open-label extension, adverse event rates were low, occurrences of LVEF <50% were uncommon, and observed reductions in LVEF were transient and effectively managed using protocol-guided dose reduction without the need for treatment interruption. Importantly, there were no cases of clinical heart failure associated with occurrences of LVEF <50% attributable to aficamten, and no occurrences of LVEF <40%, providing reassurance regarding the long-term safety of aficamten when used with individualized dose titration and monitoring.

Within this context, the low incidence of LVEF <50% observed in this analysis is clinically meaningful. All observed LVEF reductions resolved with dose modification, except for 1 patient who also demonstrated systolic dysfunction while on placebo in the parent study. This experience underscores the rapid reversibility of the pharmacodynamic effect of aficamten, without the need for treatment interruption.

Several pharmacologic properties of aficamten may contribute to this safety profile. Aficamten has a relatively short half-life, rapid washout, low liability for drug–drug interactions, and a linear pharmacokinetic/pharmacodynamic relationship,^15^ allowing for timely adjustment of drug exposure in response to changes in ventricular function. The consistent pattern of rapid recovery following dose adjustment observed in this analysis aligns with these pharmacologic characteristics and supports the safety of flexible dosing strategies in clinical practice.

The burden and clinical utility of echocardiographic monitoring during long-term therapy with cardiac myosin inhibitors is an important consideration for both patients and clinicians. In this analysis, a large number of echocardiograms were performed during the maintenance phase, yet only a small fraction led to dose reductions due to occurrences of LVEF <50%. Furthermore, the modest fluctuations in LVEF as observed in this report may fall within the expected inter-reader variability of 2D echocardiography in HCM.^16,17^ These observations raise important questions regarding the optimal frequency and intensity of surveillance imaging once patients are established on a stable maintenance dose, particularly in the absence of symptoms or prior systolic abnormalities. While continued monitoring remains essential within the current regulatory framework, these data suggest that long-term management may be simplified in the future as real-world experience with aficamten expands.

Beyond systolic function, the rates of atrial fibrillation, syncope, stroke, myocardial infarction, and death with aficamten were low and similar to placebo or metoprolol in the controlled pool, and rates of these events were generally even lower with long-term treatment of aficamten in the cumulative pool. These findings are notable given the elevated baseline risk of arrhythmias and adverse cardiovascular events in patients with oHCM.^18^ This further supports the overall cardiovascular safety of aficamten when appropriately dosed. Cardiac myosin inhibition directly targets the hypercontractile state that underlies oHCM, offering a distinct approach compared to traditional negative inotropic agents.^10^ However, because myocardial contractility is modulated at the level of the cardiac sarcomere, excessive exposure carries an inherent risk of systolic dysfunction. This potential serves as the basis for the REMS required by the US FDA for approved agents in this class.

Reports of hypertension were more frequent in aficamten-treated patients compared with placebo or metoprolol and were consistent with small increases of blood pressure observed with aficamten treatment. However, these increases of blood pressure and reports of hypertension appear to be associated with relief of LVOT obstruction and improved hemodynamics rather than an off-target side effect of aficamten. Similar increases of blood pressure are observed with relief of LV outflow obstruction with mavacamten use, septal myectomy for oHCM, and transcatheter aortic valve implantation for aortic stenosis and are likely related to subsequent increases in stroke volume and cardiac output.^19–21^ Importantly, increases of blood pressure have not been observed with aficamten treatment in patients with non-obstructive HCM.

Overall, the results of this integrated safety analysis provide empirical support for the REMS program for aficamten. The low incidence of systolic dysfunction, absence of associated heart failure, and consistent reversibility of LVEF reductions with protocol-guided dose adjustments align with the regulatory framework that permits flexible titration without routine treatment interruption (Central Illustration).

This analysis has inherent limitations. The dataset includes trials with similar but somewhat different designs, durations, and comparators, and the open-label extension lacks a concurrent control group. In addition, while approximately 700 patient-years of data were available, even longer-term experience is desirable.

## CONCLUSIONS

This integrated safety analysis demonstrates that aficamten is well tolerated in patients with oHCM across a wide range of clinical settings and durations of exposure. Systolic dysfunction was uncommon, reversible, not associated with heart failure, and effectively managed through protocol-guided dose adjustment. Taken together with previously reported efficacy data, these findings further support a favorable benefit-risk ratio and the long-term use of aficamten as a targeted therapy for oHCM.

## CLINICAL PERSPECTIVES

### COMPETENCY IN MEDICAL KNOWLEDGE

In patients with oHCM, aficamten demonstrates a favorable long-term safety profile across randomized controlled trials, an active-comparator study, and an open-label extension. Reductions in LVEF were uncommon, reversible with protocol-guided dose adjustment, and not associated with clinical heart failure. Rates of atrial fibrillation and other major cardiovascular adverse events were low and similar to placebo or metoprolol. These findings support the safe incorporation of aficamten into long-term management strategies for oHCM when accompanied by individualized dose titration and monitoring.

### TRANSLATIONAL OUTLOOK 1

Although current regulatory requirements mandate regular echocardiographic monitoring during treatment with cardiac myosin inhibitors, this analysis demonstrates that a very large volume of surveillance echocardiograms resulted in a very small number of dose reductions during the maintenance phase. Future studies should evaluate whether monitoring strategies can be optimized or individualized after patients achieve a stable maintenance dose, balancing patient safety with the practical burden of long-term imaging.

### TRANSLATIONAL OUTLOOK 2

Pharmacologic differences between aficamten and mavacamten may influence dosing flexibility, reversibility of systolic effects, and long-term safety. Prospective comparative studies and real-world evidence will be important to further define how these differences translate into clinical outcomes, optimal monitoring paradigms, and patient selection across diverse oHCM populations.

## ACKNOWLEDGMENTS

The study team and the authors thank all patients who participated in these studies and their families. They also thank all study sites and research personnel for their contributions. Editorial support was provided by Elyse Smith, PhD, CMPP, on behalf of Engage Scientific Solutions, and was funded by Cytokinetics, Inc.

## DATA AVAILABILITY STATEMENT

Qualified researchers may submit a request containing the research objectives, end points/outcomes of interest, statistical analysis plan, data requirements, publication plan, and qualifications of the researcher(s). In general, Cytokinetics does not grant external requests for individual patient data for the purpose of reevaluating safety and efficacy issues already addressed in the product labeling. Requests are reviewed by a committee of internal advisors, and if not approved, may be further reviewed by a Data Sharing Independent Review Panel. Upon approval, information necessary to address the research question will be provided under the terms of a data sharing agreement. This may include anonymized individual patient data and/or available supporting documents, containing fragments of analysis code where provided in the analysis specifications. Requests may be submitted to.

## ABBREVIATIONS AND ACRONYMS

EAIR: exposure-adjusted incidence rate
LVEF: left ventricular ejection fraction
LVOT: left ventricular outflow tract
oHCM: obstructive hypertrophic cardiomyopathy
TEAE: treatment-emergent adverse event

---

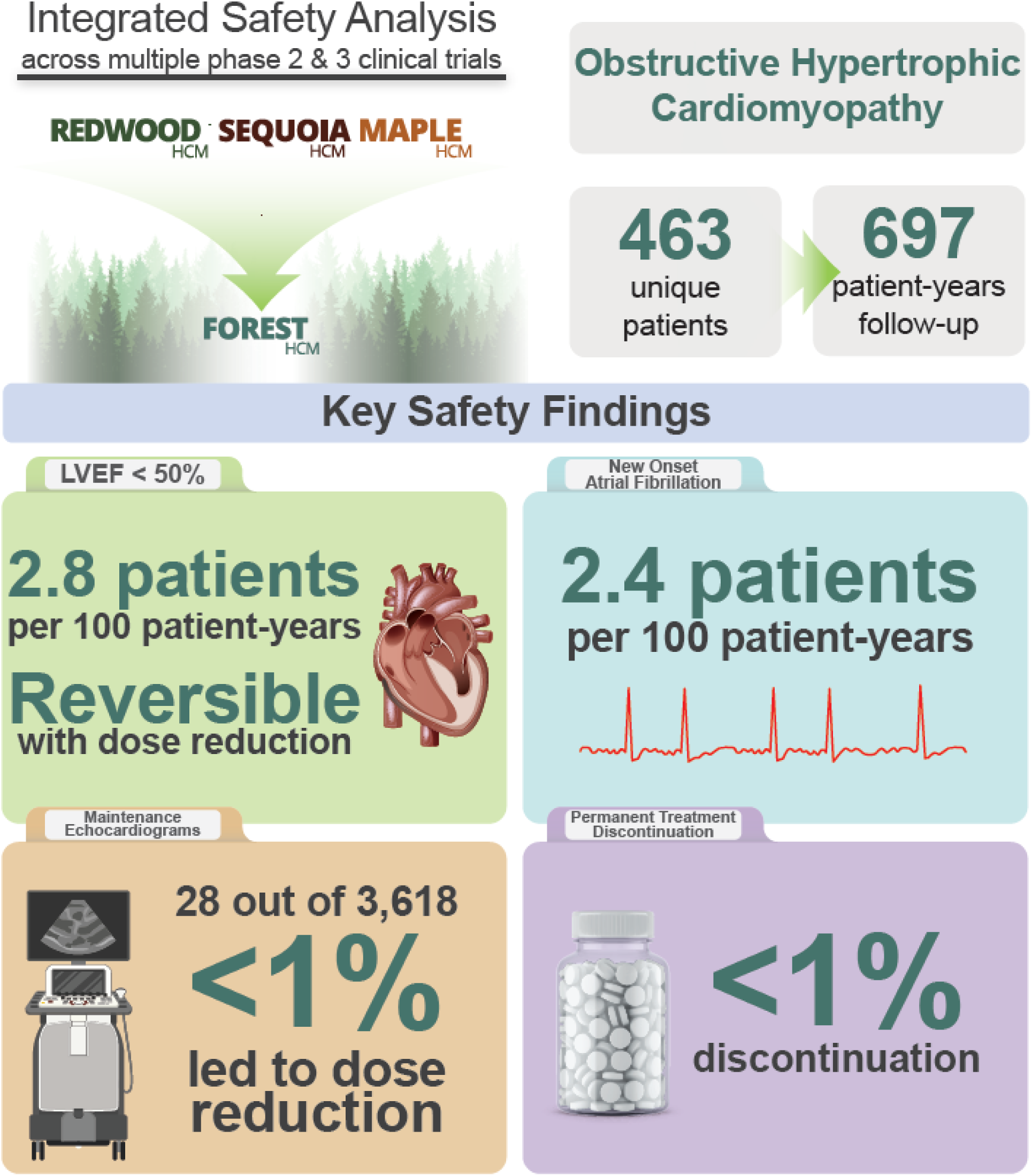
CENTRAL ILLUSTRATION – Key findings of the integrated safety analysis.

